# Two-week supplementation of *Bifidobacterium adolescentis* iVS-1 reduces fecal urgency and diarrhea and enhances overall lactose tolerance in lactose maldigesters

**DOI:** 10.1101/2024.12.16.24319107

**Authors:** Monica Ramakrishnan, Tzu-Wen L. Cross, Anne K. Wilcox, Anna Clapp Organski, Robin L. Rhine, Sindusha Mysore Saiprasad, Abigayle M. R. Simpson, Daniel J. Tancredi, Mallory J. Van Haute, Chloe M. Christensen, Zachery T. Lewis, Thomas A. Auchtung, Jens Walter, Robert Hutkins, Dennis A. Savaiano

## Abstract

Probiotic supplements containing high β-galactosidase-producing bacteria may aid in the management of lactose intolerance. We previously isolated a strain of *Bifidobacterium adolescentis*, iVS-1, from the fecal sample of a human donor after consumption of galactooligosaccharides (GOS), a prebiotic derived from lactose. Therefore, it was hypothesized that iVS-1 might reduce symptoms associated with lactose maldigestion. Compared to other probiotic strains, iVS-1 had high β-galactosidase activity and reduced gas formation by fecal communities during in vitro fermentations of lactose or milk. A randomized placebo-controlled clinical trial was then conducted with 21 lactose maldigesters, randomized to receive either *B. adolescentis* iVS-1 (n = 11) or placebo (n = 10) daily for two weeks. Compared to the two-week run-in period, iVS-1 abundance was higher at the end of the treatment period (p = 0.0005) and after the two week post-treatment period (p = 0.045). The iVS-1 group experienced less overall daily symptoms during the treatment period when compared to placebo (p = 0.032) and had significant improvement for fecal urgency (p = 0.033) and diarrhea (p = 0.006). The metabolism of lactose, reduction of gas, and improvement of multiple gastrointestinal symptoms suggests *B. adolescentis* iVS-1 may be an effective treatment for lactose intolerance.

**Trial Registration:** The trial is registered at ClinicalTrials.gov (https://clinicaltrials.gov/study/NCT05668468).

## INTRODUCTION

Lactose intolerance is a condition in which individuals experience gastrointestinal symptoms following consumption of lactose-containing products, including milk and ice cream (1). Lactose intolerance occurs in individuals who are unable to produce sufficient amounts of β-galactosidase in the small intestine, such that lactose reaches the colon and is metabolized by microbes that produce acids and gas (2). Although some lactose maldigesters can safely consume small amounts of lactose, the condition is commonly managed by eliminating lactose-containing foods from the diet (3). In contrast, yogurt, kefir, and other cultured milk products, despite containing nearly as much lactose as milk, are generally well tolerated, due to expression and in situ release of β-galactosidase by the starter culture bacteria (4).

Likewise, many studies have assessed the ability of probiotic bacteria, including specific strains of lactobacilli and bifidobacteria, to consume lactose and reduce symptoms associated with lactose maldigestion (5). In one study, two-week supplementation with *Bifidobacterium longum* capsules and *Bifidobacterium animalis* subsp. *lactis-*enriched yogurt was conducted in lactose-intolerant subjects. Both strains were detected in stool samples during supplementation (6). It was also noted that symptoms improved post-supplementation (6). There is a mixed record for improvement of symptoms due to *Bifidobacterium* supplementation in other acute and chronic studies (7-13). Some studies tested only *Bifidobacterium* strains (7, 11, 12), while the other studies included an additional probiotic with *Bifidobacterium* (8-10, 13).

In this study, we conducted a randomized clinical pilot trial to examine whether lactose intolerance symptoms could be improved by consumption of *Bifidobacterium adolescentis* iVS-1. This strain was isolated from a stool sample of a human subject after enrichment through consumption of galactooligosaccharides (GOS) and grew well, in vitro, on GOS (14, 15). Because GOS, like lactose, is hydrolyzed by β-galactosidase, we hypothesized that this autochthonous strain could also digest and consume lactose well in human subjects. Subsequent analysis of the iVS-1 genome also revealed nine β-galactosidase genes (16). Accordingly, the primary objective of this study was to assess the suitability of *B. adolescentis* iVS-1 to improve symptoms associated with lactose intolerance via both in vitro experiments and a clinical trial of lactose maldigesters given iVS-1 as a dietary supplement.

## RESULTS

### β-Galactosidase activity

*B. adolescentis* iVS-1 had been isolated from a human who consumed increasing amounts of galactooligosaccharides (GOS), a prebiotic that (similarly to lactose) is metabolized by β-galactosidase (15). Therefore, we measured the β-galactosidase activity of iVS-1 after growth on either GOS or lactose as the carbohydrate source and compared its activity with other commercial and culture collection strains of bifidobacteria and lactobacilli. When whole cells were used, iVS-1 had more activity than all other strains on GOS (146 Adjusted Miller Units; AMU) and lactose (78 AMU) (Figure 1A, Supplementary Table 1A). The type strain of *B. adolescentis*, ATCC 15703, in addition to *L. bulgaricus* ATCC 11842 and *L. acidophilus* 007, also produced measurable β-galactosidase when grown on GOS or lactose, while measurable β-galactosidase was only detected from *S. thermophilus* ATCC 19258 when grown on lactose (all between 8 - 56 AMU). The 12 other strains tested had no detectable activity under standard assay conditions. *B. adolescentis* iVS-1 also produced one of the highest amounts of β-galactosidase when the assay was conducted on lysed cells (Supplementary Figure 1).

**Figure 1.**
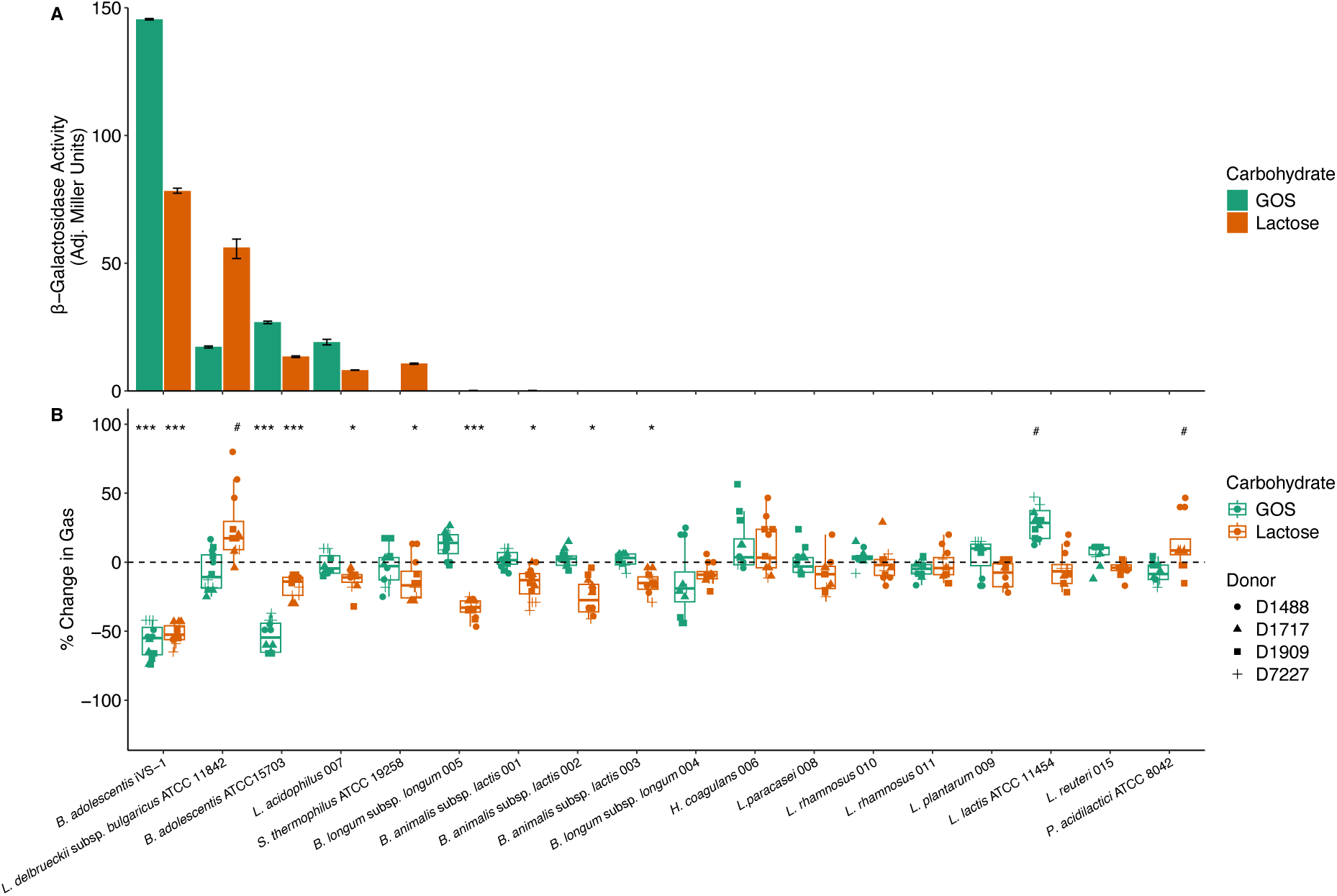
β-Galactosidase activity and gas reduction. A. Beta-galactosidase activity of whole cells grown in GOS (green) or lactose (orange) to mid-log phase, then harvested and assayed for 5 min in triplicate. Bars extend to the mean and whiskers span the 95% confidence interval. B. Change in gas produced by four fecal communities containing GOS (green) or lactose (orange), relative to communities with no probiotic added. Each shape is a different fecal donor. Box plots show the interquartile range (IQR; boxes), median (line), and 1.5 IQR (whiskers). *p < 0.05 gas reduction, ***p < 0.0005 gas reduction, #p < 0.05 gas production by Mann-Whitney-Wilcoxon test. See Supplementary Table 1 for full data and p-values. Strains are ordered by magnitude of beta-galactosidase and gas reduction, then taxonomy.

### Probiotic-mediated intestinal gas reduction

Next, we examined whether iVS-1 could reduce gas formation in an in vitro fecal fermentation model to assess the ability of the strain to compete for lactose against potential gas-forming members of the microbiota. When fecal communities were dosed with either GOS or lactose (Figure 1B), gas production was reduced when iVS-1 was added (57% reduction in GOS, 52% reduction in lactose) (Supplementary Table 1B, C). Additionally, iVS-1 reduced gas when milk was tested (32% reduction; Supplementary Figure 2). The *B. adolescentis* type strain, ATCC 15703, was also efficient at reducing gas from GOS (54%), but less so for lactose and milk (17% and 18%, respectively). Multiple other strains reduced gas production (as much as 26% reduction), but effects were smaller than seen with iVS-1. Some strains also increased gas (up to 8%).

### Clinical Trial Overview

The ability of iVS-1 to express high levels of β-galactosidase and reduce gas formation in an in vitro model suggested this strain may be effective at reducing symptoms associated with lactose intolerance. Therefore, we conducted a clinical pilot trial in participants with diagnosed lactose maldigestion. Participant characteristics are described in Table 1. There were no significant differences in the baseline characteristics of the treatment groups (Supplementary Table 1D). Twenty-one adults underwent three phases of the trial, each two weeks long (Figure 2). Subjects were to avoid milk and soft dairy products during Phases 1 and 2, then resume their normal diet during Phase 3. Phase 1 was the baseline before treatment. During Phase 2, subjects consumed a daily capsule containing either iVS-1 (n = 11 subjects) or placebo (n = 10). Phase 3 was a washout period to assess any changes post-treatment. Abdominal pain, bloating, flatulence, diarrhea, and fecal urgency were self-reported daily for the 6 weeks. Subjects were compliant in completing symptom surveys on most days, except for subject 5, who did not report any symptom scores during Phase 3. At the end of Phases 1 and 2, lactose challenges were conducted, where breath hydrogen and symptom scores were recorded over a 6 h period. In addition, the fecal bacteria composition and iVS-1 abundance were analyzed after each phase.

**Figure 2.**
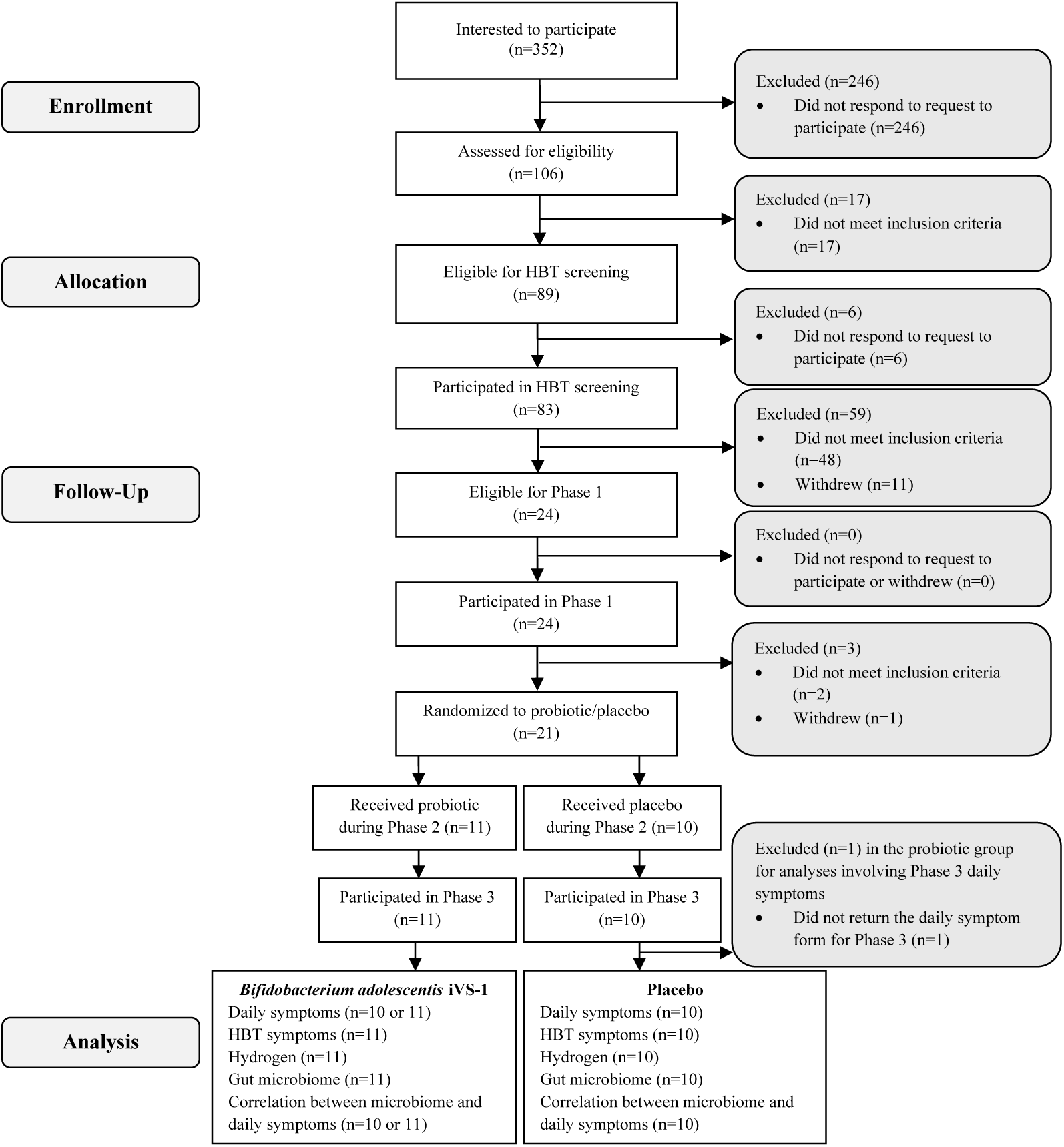
CONSORT Diagram. HBT, hydrogen breath test following lactose challenge.

**Table 1.**
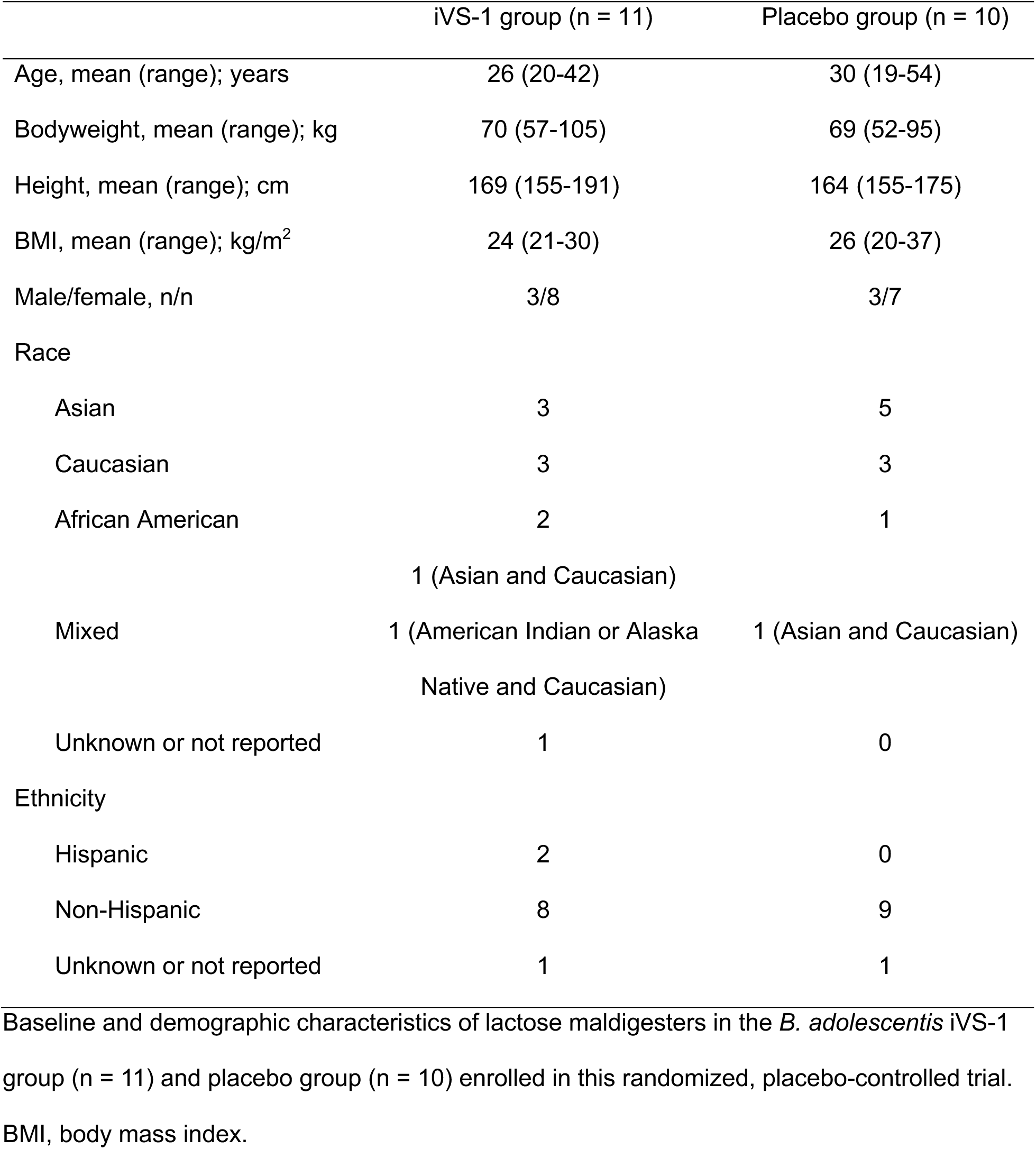
Baseline characteristics.

### Daily symptoms reported before, during, and after treatment

Mean daily scores for individual symptoms averaged together by subject and phase were consistently lower when iVS-1 was consumed (Phase 2) (Figure 3A, Supplementary Table 1E, F). The iVS-1-treated group experienced less symptoms than the placebo group during treatment: abdominal pain, 71% lower on average (p = 0.137); bloating, 53% lower (p = 0.353); diarrhea, 88% lower (p = 0.093); fecal urgency, 80% lower (p = 0.062); flatulence, 49% lower (p = 0.339). Although none of the individual comparisons was statistically significant, the composite score (sum of mean symptom scores) was significantly lower (65%; p = 0.032) for the iVS-1 group compared to the placebo during Phase 2 (Figure 3B). During Phases 1 and 3, there were no significant differences between the iVS-1 and placebo group (all p > 0.33).

**Figure 3.**
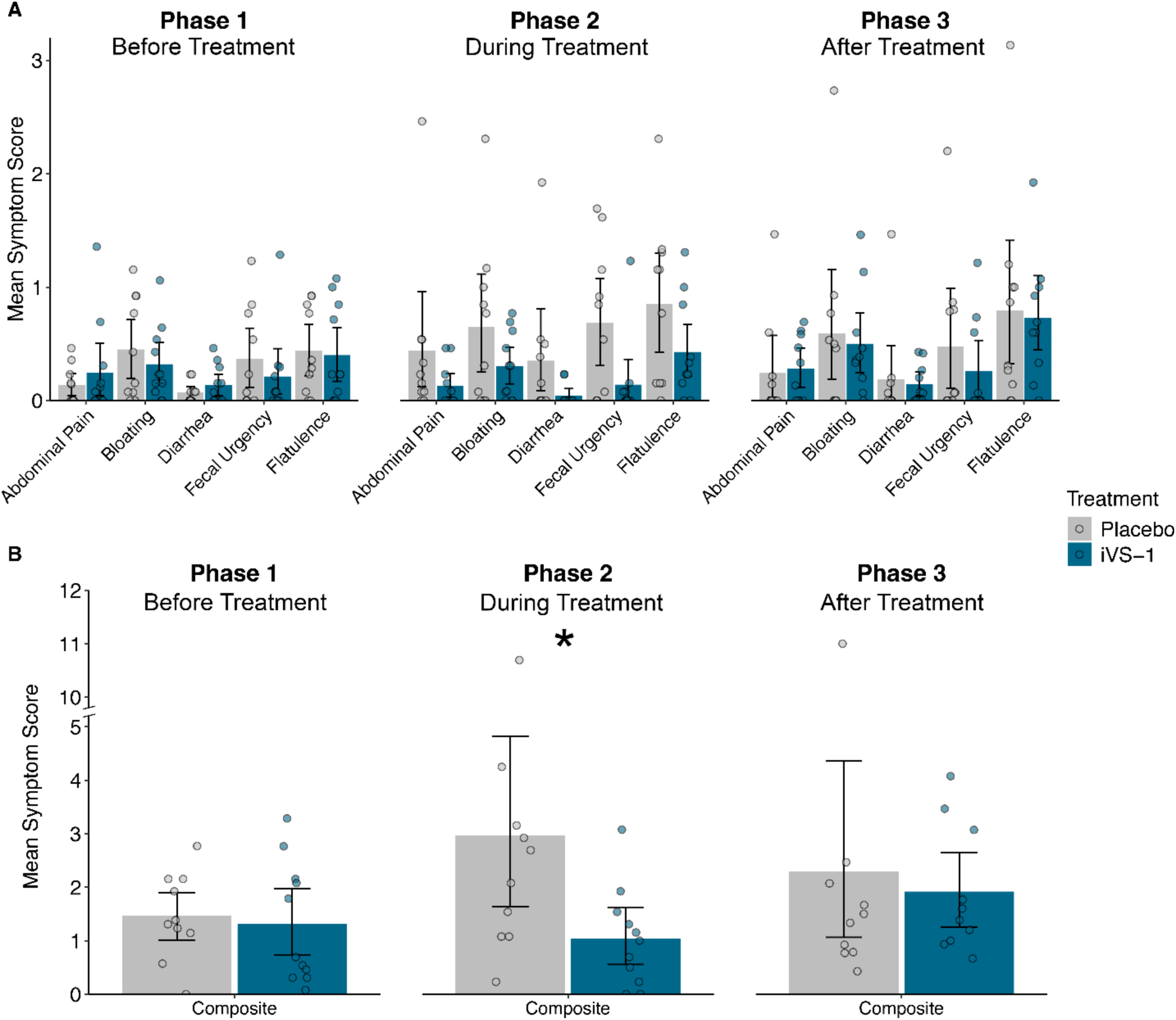
Patient daily symptom scores, by study phase. A. Mean daily symptom scores for subjects by phase. B. Composite (sum) of all mean daily symptom scores. *p < 0.05 by Mann-Whitney-Wilcoxon test. See Supplementary Table 1 for data and p-values. Bars extend to the mean and whiskers span the 95% confidence interval.

Changes in symptom scores between phases were also examined. From Phase 1 to Phase 2, mean symptom scores consistently decreased more for subjects in the iVS-1 group than the placebo group, with fecal urgency significantly lower (p = 0.046; Supplementary Figure 3A, Supplementary Table 1G, H). Following treatment (Phase 3), the composite symptom score decreased for the placebo group and increased for the iVS-1 group, and this difference was significant (p = 0.049; Supplementary Figure 3B). When individual patient daily symptom scores were the units of analysis in mixed-effects ordinal logistic regression analysis, we found that compared to the placebo group, the iVS-1 treated patients had favorable changes in the odds of reporting more severe symptoms from Phase 1 to Phase 2 for all symptoms, with the between-arm ratio of these within-person Phase 2 to Phase 1 odds-ratios being statistically significant for both fecal urgency (Odds Ratio Ratio (ORR) = 0.18 (95% Confidence Interval: 0.04 to 0.87); p = 0.033) and diarrhea (ORR = 0.03 (0.002 to 0.37); p = 0.006)(Figure 4; Supplementary Table 1I).

**Figure 4.**
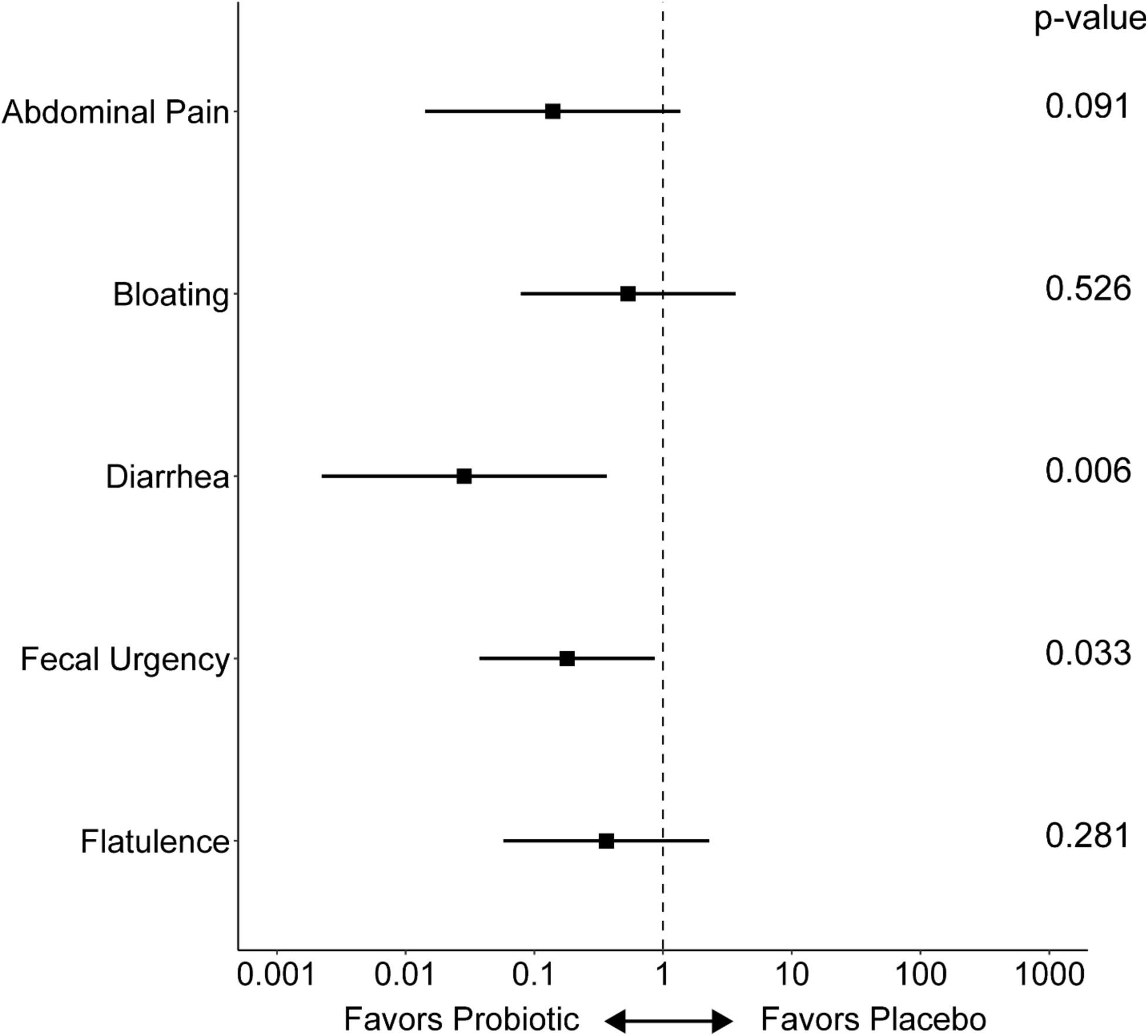
Odds ratio ratios (ORR) from mixed-effects ordinal logistic regression analysis of changes in daily symptoms from Phase 1 to Phase 2. Analyses compared the iVS-1 group with the placebo group. n = 21 subjects. See Supplementary Table 1 for all data and p-values. ORR are plotted as points, with whiskers spanning the 95% confidence interval.

### Lactose challenge breath hydrogen and symptoms

At the end of Phases 1 and 2, subjects underwent a lactose challenge where they consumed lactose water and were monitored over the following six hours for production of hydrogen gas (Supplementary Figure 4, Supplementary Table 1J-L) and symptoms (Figure 5, Supplementary Table 1M, N). For hydrogen, there were no significant differences between treatment groups (iVS-1 vs placebo) or between treatment times (post-Phase 1 vs post-Phase 2). However, among the symptom scores, fecal urgency was significantly reduced from post-Phase 1 to post-Phase 2 for the iVS-1 group (p = 0.024 by Wilcoxon signed-rank test), but not for the placebo group (p = 0.232).

**Figure 5.**
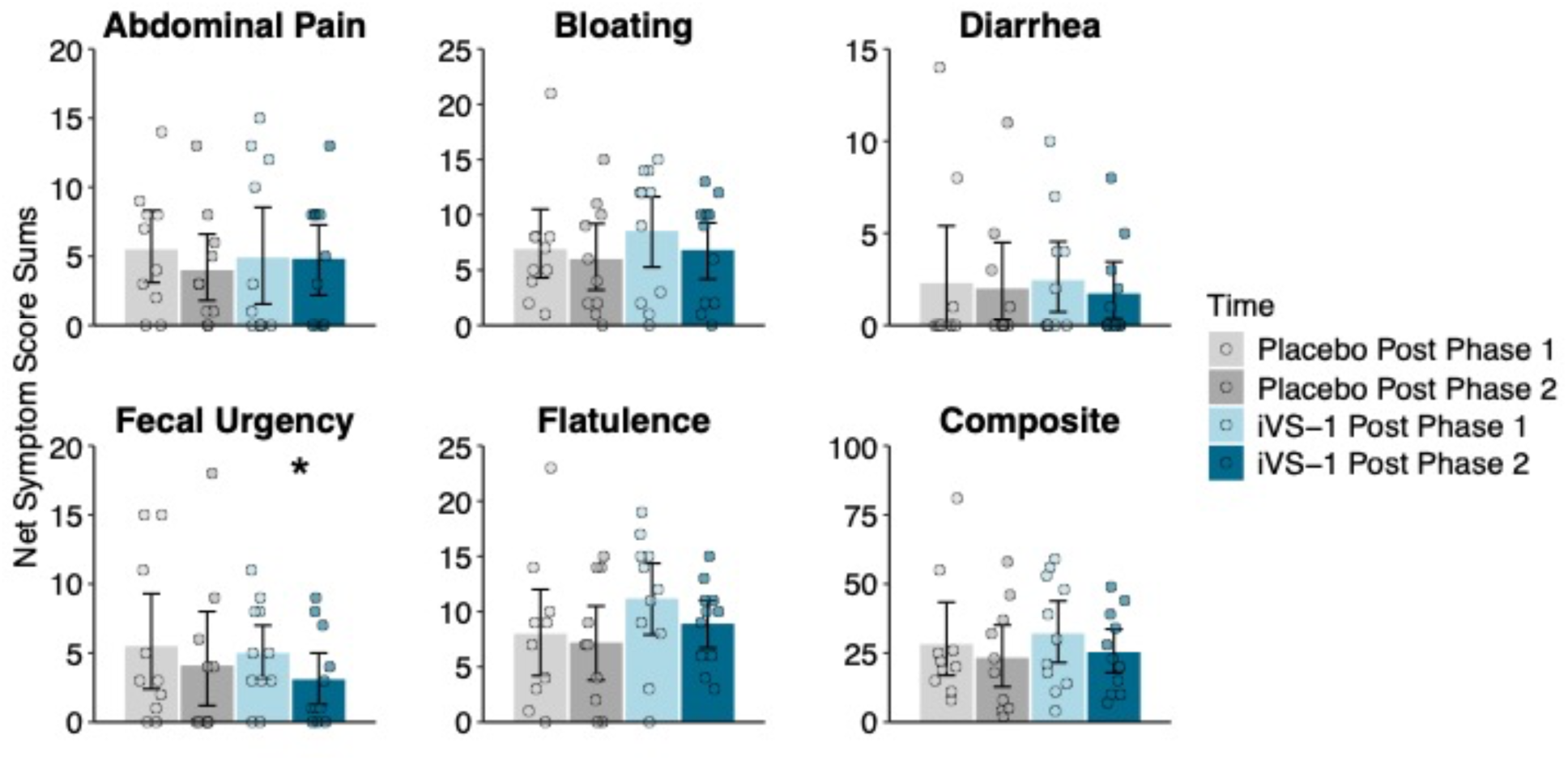
Subject symptom scores following lactose challenge. Mean symptom scores for individual symptoms and composite (sum) of all five mean symptom scores. Each subjects’ data was normalized for background symptom scores at time zero. Bars extend to the mean and whiskers span the 95% confidence interval. n = 21 subjects.*p < 0.05 by Wilcoxon signed-rank testing. Post-Phase 1 occurred between Phase 1 (before treatment) and Phase 2 (during treatment). Post-Phase 2 occurred between Phase 2 (during treatment) and Phase 3 (after treatment). See Supplementary Table 1 for data and p-values.

### Gut Microbiome

The bacterial 16S rRNA gene diversity and composition was examined in fecal samples collected following each phase. There were no significant differences in alpha or beta diversity between phases or treatment groups (Supplementary Figure 5), and no significant taxonomic differences (all FDR-adjusted p > 0.05). Since iVS-1 treatment has previously been associated with a bifidogenic effect (17), we examined the relative abundance of *Bifidobacterium* (Supplementary Figure 6, Supplementary Table 1O, P). Though not significantly different between phases or treatment groups, *Bifidobacterium* appeared to reach higher relative abundance in the iVS-1 group (6.9%) than the placebo group (5.6%) at the end of Phase 2 (p = 0.223). From the end of Phase 1 to the end of Phase 2, there was also a 0.8% increase in *Bifidobacterium* in the iVS-1 group and a 3.5% decrease in the placebo group (p = 0.085).

To examine establishment and persistence of iVS-1, strain-specific quantitative PCR was used. While iVS-1 abundance was not significantly higher in the iVS-1 group at baseline (end of Phase 1; detected in 2/11 subjects, median at the limit of detection of 500 CFU/ng DNA, mean 7 x 10^6^ CFU/ng DNA, p = 0.189), abundance was significantly higher after treatment (end of Phase 2; detected in 9/11 subjects, median 5 x 10^5^ CFU/ng DNA, mean 1 x 10^7^ CFU/ng DNA, p = 0.0005) and after the washout period (end of Phase 3; detected in 4/11 subjects, median 500 CFU/ng DNA, mean 1 x 10^7^ CFU/ng DNA, p = 0.045) (Supplementary Figure 7, Supplementary Table 1Q). Throughout the study, no samples from the placebo group had detectable iVS-1.

We next examined whether there were correlations between either the relative abundance of *Bifidobacterium* or the qPCR-determined absolute abundance of iVS-1 with any of the symptoms that occurred in that phase. While there were no significant correlations with iVS-1, significant negative correlations between the relative abundance of *Bifidobacterium* and daily bloating scores were observed during Phase 1 (R = -0.47, p = 0.038), Phase 3 (R = -0.47, p = 0.035), and all three phases combined (R = -0.46, p = 0.0002; Supplementary Figure 8 Supplementary Table 1R-T).

## DISCUSSION

Most of the world is lactose intolerant due to age-related downregulation of the β-galactosidase gene (2, 18). Although incidence varies depending on ethnicity and family genetics, about 60% of adults do not produce sufficient β-galactosidase to adequately digest lactose (2). However, studies suggest that even those individuals who cannot make the enzyme can still consume lactose without ill effect provided they contain enough β-galactosidase-producing bifidobacteria in their gastrointestinal tract (18, 19). In addition, *B. adolescentis* is overrepresented in the gut microbiome of Mongolians, an ethnic group that is largely lactose intolerant but nevertheless consumes a high amount of dairy products (20).

We hypothesized that providing a high β-galactosidase-producing *B. adolescentis* supplement to lactose maldigesters would reduce symptoms associated with lactose intolerance. Our in vitro tests showed the suitability of *B. adolescentis* iVS-1 as a candidate strain for this purpose, as it had higher β-galactosidase activity and significantly reduced gas production by members of the fecal microbiota in the presence of lactose when compared to other strains. The gas production assay confirmed an important component of the desired mechanism of action which had been missing from the screening and selection process of previous lactose maldigestion probiotic candidates. The simple presence of high β-galactosidase activity may not be sufficient for optimal symptom relief if the lactose is not diverted away from gas-producing members of the native fecal microbiota. The ability to compete against other microbes for the carbohydrate of interest in a mixed community is therefore essential. As *B. adolescentis* iVS-1 had been isolated following in vivo selection under competitive conditions and was autochthonous to the human gut (15), this strain was predicted to improve symptoms associated with lactose maldigestion under competitive conditions.

Consistent with this prediction, in the clinical trial, the iVS-1 group experienced a significant improvement in the composite symptom score during the treatment phase of the clinical trial compared to the placebo group. Additionally, when changes in symptom scores between Phase 1 (pre-treatment) and Phase 2 (post-treatment) were compared, significantly improved scores for fecal urgency and diarrhea were observed. Fecal urgency is a common symptom for people experiencing gastrointestinal symptoms, including constipation and diarrhea (21), including those with lactose intolerance (22). This study is consistent with others that have demonstrated improved lactose intolerance symptoms following supplementation with probiotics (6, 7, 10, 23). While symptoms significantly improved during iVS-1 treatment, all symptoms appeared to worsen after the treatment ended (Phase 2 to Phase 3), with a significant difference in the composite score between iVS-1 and placebo groups.

Study participants were instructed to eliminate dairy products from their diet during the run-in (Phase 1) and while they were consuming placebo or iVS-1 (Phase 2). Still, individual and composite symptoms scores during Phase 2 among the placebo group were generally about double that during Phase 1. In contrast, for the iVS-1 group, symptom scores remained the same or decreased in Phase 2. Although it is possible iVS-1 improved symptoms independent of lactose maldigestion, participants had been informed that the trial was designed to assess changes when consuming dairy. Thus, they may not have been fully compliant with the instructions to continue avoiding dairy during treatment.

The results did not reveal a significant difference in breath hydrogen levels following the two-week iVS-1 supplementation. The utility of the HBT, although long used as a diagnostic tool for lactose intolerance, has recently been questioned (24), with the authors advocating that symptoms of lactose intolerance are more meaningful. Although another recent study showed that *Bifidobacterium animalis* subsp. *lactis* Bi-07 reduced breath hydrogen compared to placebo, doses were exceptionally high at 2 × 10^12^ CFUs (11).

While supplementation affected iVS-1 absolute abundance during and after treatment, it did not lead to significant alterations in taxa, as measured by 16S rRNA relative abundance. However, levels of the genus *Bifidobacterium* were higher after treatment, consistent with a previous study, where iVS-1 enriched for *Bifidobacterium* (25). In the current study, we also observed a significant correlation between *Bifidobacterium* abundance and reduced bloating. This observation is consistent with other studies showing high abundance of *Bifidobacterium*, including *B. adolescentis*, is associated with lactose tolerance among lactase nonpersistent individuals (18, 19).

## CONCLUSIONS

In this study, we established that *B. adolescentis* iVS-1 has high β-galactosidase activity and can reduce gas formation from lactose during in vitro fermentations. In a pilot clinical trial, we provide evidence that supplementation with this strain can provide benefits to lactose maldigesters for managing symptoms, including fecal urgency and diarrhea. Future studies with more participants are encouraged to further support these findings.

## METHODS

### β-Galactosidase assays

β-Galactosidase assays were conducted using a protocol modified from that of Miller (26). Cells were prepared by inoculation of fresh colonies into reduced MRS broth (BD Difco) supplemented with 0.05% cysteine, followed by anaerobic incubation at 37°C for 12 h. One hundred microliters of the grown culture was transferred into each of two tubes of 9.9 mL of reduced Complex Gut Media + MOPS (CGMM), a media formulated to simulate gastrointestinal conditions. CGMM is composed of peptone (0.5 g/L), yeast extract (1 g/L), sodium chloride (1 g/L), magnesium sulfate (0.1 g/L), calcium chloride (0.1 g/L), hemin (.005 g/L), cysteine (0.5 g/L), bile salts (0.5 g/L), sodium acetate (1.35 g/L), sodium propionate (0.43 g/L), isobutyric acid (0.5 mM), isovaleric acid (0.5 mM), valeric acid (0.5 mM), sodium bicarbonate (4 g/L), potassium phosphate (0.45 g/L each of KH2PO4 and K2HPO4), vitamin K1 (1 mg/L), maltose (0.1 g/L), cellobiose (0.1 g/L), inulin (0.1 g/L), arabinogalactan (0.1 g/L), soluble starch (0.1 g/L), and 3-(N-morpholino)propanesulfonic acid (MOPS) buffer pH 7.3 (100 mM). Here, CGMM was supplemented with either 1% lactose (100% purity, Loudwolf) or galactooligosaccharides (GOS; 95% purity, Shanghai Freemen). Cells were grown at 37°C in for collection at 6 h. Cells were harvested by centrifugation and the pellet was resuspended in 10 mL of Z buffer (disodium phosphate, 16.1 g/L; monosodium phosphate, 5.5 g/L; potassium chloride, 0.75 g/L; magnesium sulfate 0.24 g/L, adjusted to pH 7.0). The cells were then centrifuged again, and resuspended in 2.5 mL Z buffer, generating a 4X concentrated cell suspension.

For whole cell assays, cell suspensions (0.25 mL) were added to tubes containing 0.75 mL Z buffer. One of these tubes was set aside for enumeration. To the remaining tube, 0.1 mL of ortho-Nitrophenyl-β-galactoside (ONPG) solution (10 mM ONPG suspended in 0.03 M sodium phosphate buffer, pH 6.8) was added. After 5 min, the reactions were centrifuged 3 min, supernatants (150 μL) were aliquoted in triplicate into 96-well plates, and 50 μL of stop solution (1 M sodium carbonate) was added. Optical densities were determined at 420 nm and 550 nm in a microplate reader (BioTek Synergy H1).

Intracellular β-galactosidase activity was also measured using lysed cells. A 1.0 mL portion of concentrated cell suspension was aliquoted into tubes containing 200 mg of 0.1 glass beads (BioSpec) and bead-beaten twice at 1800 RPM for 1 min intervals, interspersed with a 1 min incubation on ice. Tubes were centrifuged and the supernatant was added 1:4 into a tube containing Z buffer. To this tube, ONPG solution was added β-galactosidase activity was measured as above. For both assays, β-galactosidase activity (modified Miller’s units) was determined using the formula 1000 × (A420 nm - 1.75 × A500 nm)/(time (min) × vol (mL) × CFU correction factor (normalized to 10^7^)), where CFU was used for a more precise measurement of cell numbers than the traditional 620 nm reading.

### Gas reduction experiments

Gas experiments were performed by incubating test strains in reduced CGMM containing lactose, GOS, or milk, with fecal samples from one of four self-described healthy donors with no gastrointestinal disorders. The four representative fecal samples were chosen based on variation in gas production in response to assorted compounds. Fecal donations were collected under Advarra IRB# Pro00059566. Individual strains were prepared by growing in Reinforced Clostridial Medium (RCM)( BD Difco recipe, except with no agar) for all strains except *Lactobacillus acidophilus* 007, which was grown on MRS. Cells were collected in late log phase, pelleted by centrifugation, resuspended in CGMM containing 7% dimethyl sulfoxide (DMSO) cryoprotectant, and frozen at -80 C. To start the experiment, cells were thawed, added to 2 mL glass vials with crew caps containing septa composed of PTFE and silicone. Cells were added at equal cell concentrations (10^7^ CFU for lactose experiment, 10^8^ CFU for GOS and milk experiments). Fecal communities (0.5 mL) were added after diluting frozen fecal samples 1:50 (lactose experiment) or 1:500 (GOS and milk experiments) in CGMM + lactose, GOS, or 2% ultra pasteurized milk (1%,1%, 5% final concentrations by volume). Experiments were conducted in an anaerobic chamber with 90% N2, 5% CO2, and 5% H2 atmosphere. After 18 h at 37°C, shaking at 125 RPM, the amount of gas was measured by inserting a needle attached to a glass syringe through the septum. Gas reduction was calculated by the formula 100 × ((mean of no probiotic control + DMSO) - probiotic test sample)/(mean of no probiotic control + DMSO). DMSO, added at different levels due to the cell suspensions being different concentrations, did not have a significant effect on gas production (Supplementary Table 1U).

### Strains used in in vitro experiments

The following strains were used in β-galactosidase assays and/or gas experiments (obtained from the following sources): *Bifidobacterium adolescentis* iVS-1 (Synbiotic Health), *Bifidobacterium adolescentis* ATCC15703, *Bifidobacterium longum* subsp. *infantis* ATCC 15697, *Heyndrickxia coagulans* 014, and *Lactococcus lactis* ATCC 11454 (academic labs), *Pediococcus acidilactici* ATCC 8042 (USDA ARS NRRL), and *Lactobacillus delbrueckii* subsp. *bulgaricus* ATCC 11842 (American Type Culture Collection). The following strains were isolated from commercial products and de-identified: *Bifidobacterium animalis* subsp. *lactis* 001, *Bifidobacterium animalis* subsp. *lactis* 002, *Bifidobacterium animalis* subsp. *lactis* 003, *Bifidobacterium longum* subsp. *longum*004, *Bifidobacterium longum* subsp. *longum 005, Heyndrickxia coagulans* 006, *Lactobacillus acidophilus* 007, *Lactobacillus paracasei* 008*, Lactobacillus plantarum 009*, *Lactobacillus rhamnosus* 010, *Lactobacillus rhamnosus 011*, *Bifidobacterium bifidum 012*, *Lactobacillus gasseri* 013, *Lactobacillus reuteri* 015, and *Streptococcus thermophilus* ATCC 19258.

### Clinical Trial Participant Recruitment and Eligibility Criteria

Twenty-one verified lactose maldigesters were enrolled in the study. Sample sizes were calculated based on a previous similarly-designed study (27). This was a randomized, placebo-controlled trial. Subjects were recruited through flyers, Purdue campus emails, and advertisements in West Lafayette, IN, USA, and campus newspapers (Purdue University, West Lafayette, IN, USA). Recruitment began in February 2022 and the study was completed in December 2023.

A total of 352 individuals expressed interest in the study and contacted the study staff via email or telephone. Of these, 89 individuals met the eligibility criteria during the phone screening (Appendix A). Eligible participants were assigned a subject identification number after they read and signed the informed consent form. Sixty-eight subjects underwent a hydrogen breath screening to verify lactose maldigestion. Twenty-one verified lactose maldigesters completed all phases of the study, except for subject 5, which did not complete daily symptom surveys for Phase 3. Out of the twenty-one subjects, eleven were randomized to the *Bifidobacterium adolescentis* iVS-1 probiotic group and ten were randomized to the placebo group. Randomization of subjects was performed by an independent organization. Participants and study personnel were blinded to the treatment; hence this was a double-blinded trial.

Eligibility criteria required participants to be aged 18 to 65, provide informed consent, and self-report lactose intolerance or maldigestion. They had to agree to refrain from using any treatments or products for dairy intolerance during the study, be willing to attend all study visits and complete all procedures, including fasting for hydrogen breath tests and avoiding lactose in their diets for part of the study. Additionally, participants needed to understand and provide written informed consent in English. Lactose maldigesters were verified with a 20 ppm increase in hydrogen levels during a three-hour hydrogen breath test (HBT) after a challenge dose of milk (Kroger® 2% reduced fat; The Kroger Co., Indianapolis, IN, USA) containing 0.5 g of lactose per kg of body weight. These verified lactose maldigesters were enrolled in the study.

During the study, subjects rated abdominal pain, bloating, flatulence, diarrhea, and fecal urgency on a six-point Likert scale from 0 (no symptoms) to 5 (severe symptoms), with intermediate values (1 for slight, 2 for mild, 3 for moderate, and 4 for moderately severe) indicating increasing severity. Stool samples were collected for gut microbiota analysis.

### Clinical Trial Exclusion Criteria

Exclusion criteria included milk allergy, current pregnancy or lactation, tobacco or nicotine use within three months, and significant gastrointestinal or systemic conditions such as gastroparesis, amyloidosis, neuromuscular diseases, and diabetes. Individuals with a history of gastrointestinal surgeries, chronic pancreatitis, inflammatory bowel diseases (IBD), active or severe ulcers, HIV, or hepatitis B or C were excluded. Recent bowel preparation, use of dairy intolerance therapies, chronic antacid or proton pump inhibitors (PPI) use, recent antibiotics (usage within 30 days prior to screening), or high colonic enemas within 30 days, alcohol or drug abuse in the past 12 months, ongoing chemotherapy, recent participation in other studies, or any other conditions impacting participation were also grounds for exclusion.

### Clinical Trial Intervention Types

There were two interventions: a placebo capsule containing microcrystalline cellulose (MCC; Comprecel M112, Mingtai Chemical, Mountainside, NJ, USA), and a probiotic capsule consisting of *Bifidobacterium adolescentis* iVS-1 (Synbiotic Health, Lincoln, NE, USA) blended with MCC. Probiotic capsules were confirmed to contain ≥10^9^ CFU through the end of the study. All capsules were identical in size, shape, and color.

### Clinical Trial Ethics

The study was registered with ClinicalTrials.gov under the identifier NCT05668468 and approved by the Purdue Institutional Review Board (IRB-2021-1099). The study adhered to the principles of the Helsinki Declaration of 1975, as revised in 2008, and followed the International Conference on Harmonization Good Clinical Practice guidelines.

### Clinical Trial Procedure

The study was conducted in three phases (Figure 2). During the first phase of the study, between days 1 and 14, participants avoided lactose intake, including milk in any form and fluid or soft dairy foods. They rated any symptoms associated with lactose intolerance once daily on a Likert scale and recorded them on the symptoms form. Participants provided a stool sample between days 11 and 13 using a stool sample collection kit following provided instructions used in the Manual of Procedures for Human Microbiome Project, Version 12.0 (28). Participants stored collected samples in a NanoCool box (Albuquerque, NM) with two ice packs to keep their samples cold. Participants were instructed to return their stool samples within 24 h of collection. Upon arrival, an aliquot of feces was collected for DNA extraction and immediately stored at −80 °C until further processing. On day 14, participants underwent a six-hour HBT, involving the consumption of water containing 0.5 g of lactose per kg of body weight followed by breath sample collection at 0, 0.5, 1, 2, 3, 4, 5, and 6 h timepoints. Participants fasted for 12 h before the HBT, consuming only water during this period and adhering to a low-fiber meal the night before. Participants with an increase of at least 20 ppm hydrogen during the HBT entered Phase 2 (days 15-27), where they continued to avoid dairy and were randomized to receive either a capsule containing *Bifidobacterium adolescentis* iVS-1 or placebo daily, continuing to record daily symptoms and providing another stool sample between days 25 and 27 using the same method described above. On day 28, participants underwent a six-hour HBT with the same lactose dose as in Phase 1. The participants were instructed to resume their habitual diet during Phase 3 (days 29-43), including dairy, and continue to record symptoms daily and provide a final stool sample between days 40 and 42. Symptoms were recorded during each 6 h HBT, and microbiota analysis was conducted on the stool samples.

### Clinical Trial Adverse Events

No adverse events were reported by any of the participants during the study.

### Gut Microbiota Analysis

Total DNA from the collected human stool samples were extracted using QIAamp PowerFecal Pro DNA Kit (Qiagen Sciences Inc., Germantown, MD) per manufacturer’s protocol. The concentration of extracted genomic DNA was quantified using a Qubit Flex Fluorometer (Life Technologies, Grand Island, NY). Amplification of 16S rRNA genes was generated from genomic DNA through PCR using unique 8-bp barcodes attached to the forward and reverse universal primers flanking the hypervariable 4 (V4) region of the bacterial 16S rRNA gene and fused with Illumina sequencing adapters (29). Each sample was amplified in duplicate in a reaction volume of 12.5 μL using KAPA HiFi HotStart DNA polymerase (KAPA Biosystems, Wilmington, MA), 10 μM of each primer, and 25 ng of genomic DNA. PCR was carried out under the following conditions: initial denaturation for 3 min at 95 °C, followed by 25 cycles of denaturation for 30 s at 95 °C, annealing for 30 s at 55 °C and elongation for 30 s at 72 °C, and a final elongation step for 5 min at 72 °C. PCR products were purified with the QIAquick 96-well PCR Purification Kit (Qiagen Sciences Inc., Germantown, MD) and then quantified using Qubit dsDNA BR Assay kit (Invitrogen, Oregon, USA). Samples were equimolar pooled and sequenced at the Genomics and Gene Editing Facility of Bindley Bioscience Center at Purdue University using the v2 chemistry of the Illumina MiSeq platform (Illumina, San Diego, CA) to generate 2x250 bp pair-end reads.

Sequencing reads were processed using the Quantitative Insights into Microbial Ecology (QIIME) 2 pipeline (2024.5) (30). Briefly, demultiplexed paired-end sequences were imported using Casava 1.8 format and denoised using DADA2 (31) to obtain an amplicon sequence variant (ASV) table. ASV present less than 10 times per sample and in less than four samples were discarded. A naive Bayes taxonomy classifier was trained on the SILVA vs. 138 (32) reference database (clustered at 99% similarity) and used to assign taxonomy to the resulting 167 ASVs (33). An even sampling depth (sequences per sample) of 10622 sequences per sample was used for assessing alpha- and beta-diversity measures. Beta diversity was assessed using principal coordinates analysis (PCoA) of unweighted and weighted UniFrac, Bray-Curtis, and Jaccard distance metrics. Alpha diversity was assessed using observed ASVs, Shannon indices, and evenness. Distances between treatment groups at each timepoint were tested by pairwise PERMANOVA (34).

### Quantitative Analysis of *Bifidobacteria adolescentis* iVS-1

Quantitative PCR (qPCR) was used to quantify *Bifidobacterium adolescentis* iVS-1 in fecal samples using the same genomic DNA used for 16S rRNA gene sequencing. Primers iVS-1-F (TTGCTTTTGCTCTGGAACATAC) and iVS-1-R (GTAATGAGGTAATACTGCGTCC) (15) were confirmed to be specific (no other close hits by BLAST; (35). Ten ng of DNA were used for each qPCR reaction, performed using PowerUp SYBR Green Master Mix (Applied Biosystems, Waltham, MA) on a QuantStudio 7 Flex Real-Time PCR system (Applied Biosystems, Waltham, MA). Amplification was executed in the following conditions: UDG activation step for 50 °C for 2 min, DNA polymerase activation at 95 °C for 2 min, followed by 40 cycles of denaturation at 95 °C for 1 s and annealing/extension at 61 °C for 30 s. Dissociation curve conditions were performed as follows: 95 °C for 15 s, 60 °C for 1 min, and 95°C for 15 s. Presence in feces was quantified against a standard curve made with pure culture of *B. adolescentis* iVS-1 that was grown in reinforced clostridial media (RCM) for 14 h at 37 °C. Bacterial culture was then quantified by serial dilution onto RCM agar plates to determine colony forming units per mL of culture (CFU/mL). DNA was isolated from the primary dilution and then was serially diluted ten-fold to create seven standards.

### Statistical Analysis

Given the modest number of patients and the skewed outcome distributions, we avoided comparisons relying heavily on normal distribution assumptions. Mann-Whitney-Wilcoxon and Wilcoxon signed-rank testing was performed in R with package ‘wilcox.test’ to examine differences in gas reduction, participant baseline characteristics, daily symptoms, hydrogen levels and symptoms during the hydrogen breath test, *Bifidobacterium* relative abundance, and/or iVS-1 abundance. For the primary analysis of symptoms, we averaged daily scores together by patient and study phase. For participants with >13 days in Phase 1 (up to 4 extra days) or Phase 3 (up to 2 extra days), symptom scores for all days were included in analyses. One participant did not submit their daily symptom forms for Phase 3 of the study. Therefore, the data for this participant were not included in the Phase 3 daily symptoms analysis. For hydrogen levels, the sum of gas measured over 6 hr from each subjects for each post study phase was analyzed. For symptoms during the HBT, we analyzed the mean, sum, max, and area under the curve for scores, by patient, symptom, and post study phase. Generalized estimating equation modeling was performed on HBT hydrogen and symptom time courses, in R using the package ‘geepack’ with a Poisson distribution and AR(1) correlation structure. We also used Stata to explore the use of mixed-effects ordinal logistic regression analysis on the patient daily scores, an approach that avoided treating intervals among adjacent levels in the response scale as equal, while also permitting within-person correlations in responses to be modeled. Taxonomic differences were assessed in ATIMA (https://atima.research.bcm.edu) with false discovery rate-adjusted Mann-Whitney-Wilcoxon p-values. Spearman correlation of *Bifidobacterium* relative abundance and iVS-1 abundance with symptoms during the two week-treatment was analyzed in R using the ‘corrplot’ package.

## ACKNOWLEDGEMENTS

A microbial strain used in this work was provided by the USDA-ARS Culture Collection (NRRL). We thank Anissa Armet for analytical advice and Car Reen Kok for strains. The graphical abstract was created with BioRender.

## DATA AVAILABILITY

16S rRNA gene data has been deposited in NCBI’s Sequence Read Archive under BioProject ID PRJNA1162892. R code used to generate figures and calculate statistics, and related files are available at https://github.com/auchtung/Ramakrishnan2024. [To be posted upon acceptance of manuscript]

## Appendix A

Inclusion Criteria:

1. Ability/desire to provide informed consent
2. Aged 18 to 65 years of age inclusive at screening
3. Self-report of lactose intolerance or lactose maldigestion
4. Agrees to refrain from all other treatments and products used for dairy intolerance (e.g., Lactaid®, Dietary Supplements, probiotics) during study involvement
5. Willing to return for all study visits and complete all study related procedures, including fasting before and during the hydrogen breath tests, and avoiding lactose in their diets (ex. milk, fluid, and soft dairy foods) for Days 1-28 of the study
6. Able to understand and provide written informed consent in English

Exclusion Criteria:

1. Allergic to milk
2. Currently pregnant
3. Currently lactating
4. Cigarette smoking or other use of tobacco or nicotine containing products within 3 months of screening
5. Diagnosed with any of the following disorders known to be associated with abnormal gastrointestinal motility such as Gastroparesis, amyloidosis, neuromuscular diseases (including Parkinson’s disease), collagen vascular diseases, alcoholism, uremia, malnutrition, or untreated hypothyroidism
6. History of surgery that alters the normal function of the gastrointestinal tract including, but not limited to gastrointestinal bypass surgery, bariatric surgery, gastric banding, vagotomy, fundoplication, pyloroplasty [Note: history of uncomplicated abdominal surgeries such as removal of an appendix more than 12 months prior to screening will not be excluded]
7. Past or present: Organ transplant, chronic pancreatitis, pancreatic insufficiency, symptomatic biliary disease, Celiac disease, chronic constipation, diverticulosis, inflammatory bowel disease (IBD), ulcerative colitis (UC), Crohn’s disease (CD), small intestine bacterial overgrowth syndrome (SIBO), gastroparesis, gastro-esophageal reflux disease (GERD), Irritable Bowel Syndrome (IBS) or any other medical condition with symptoms that could confound collection of adverse events.
8. Active ulcers, or history of severe ulcers
9. Diabetes mellitus (type 1 and type 2)
10. Congestive Heart Failure (CHF)
11. Human Immunodeficiency Virus (HIV), Hepatitis B or Hepatitis C
12. Recent bowel preparation for endoscopic or radiologic investigation within four weeks of screening (e.g., colonoscopy prep)
13. Use of concurrent therapy(ies) or other products (e.g., laxatives, stool softeners, Pepto Bismol®, Lactaid® Dietary Supplements, probiotics) used for symptoms of dairy intolerance within 7 days of screening
14. Chronic antacid and/or PPI use
15. Recent use of systemic antibiotics defined as use within 30 days prior to screening
16. Recent high colonic enema, defined as use within 30 days prior to screening
17. Any concurrent disease or symptoms which may interfere with the assessment of the cardinal symptoms of dairy intolerance (i.e., gas, diarrhea, bloating, cramps /stomach pain, fecal urgency)
18. History of ethanol (alcohol) and/or drug abuse in the past 12 months
19. Currently undergoing chemotherapy
20. Use of any investigational drug or participation in any investigational study within 30 days prior to screening
21. Prior enrollment in this study
22. Any other conditions/issues noted by the study staff and/or Principal Investigator that would impact participation and/or protocol compliance

## SUPPLEMENTARY FIGURES

**Supplementary Figure 1.**
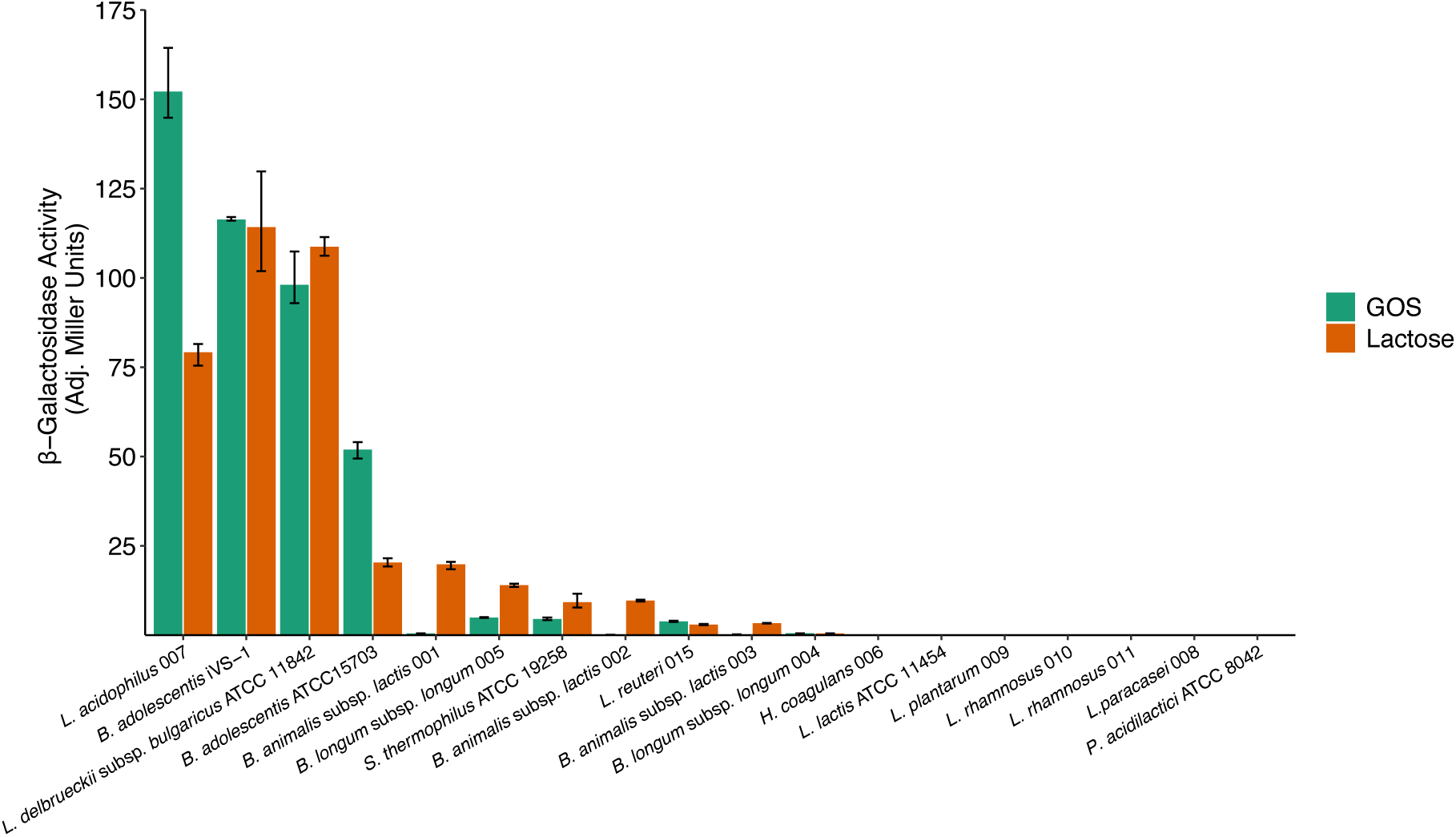
β-galactosidase activity of lysed cells. β-galactosidase activity of cells grown in GOS (green) or lactose (orange) to mid-log phase, then harvested, lysed, and assayed for 5 min. See Supplementary Table 1 for data and p-values. Bars extend to the mean and whiskers span the 95% confidence interval.

**Supplementary Figure 2.**
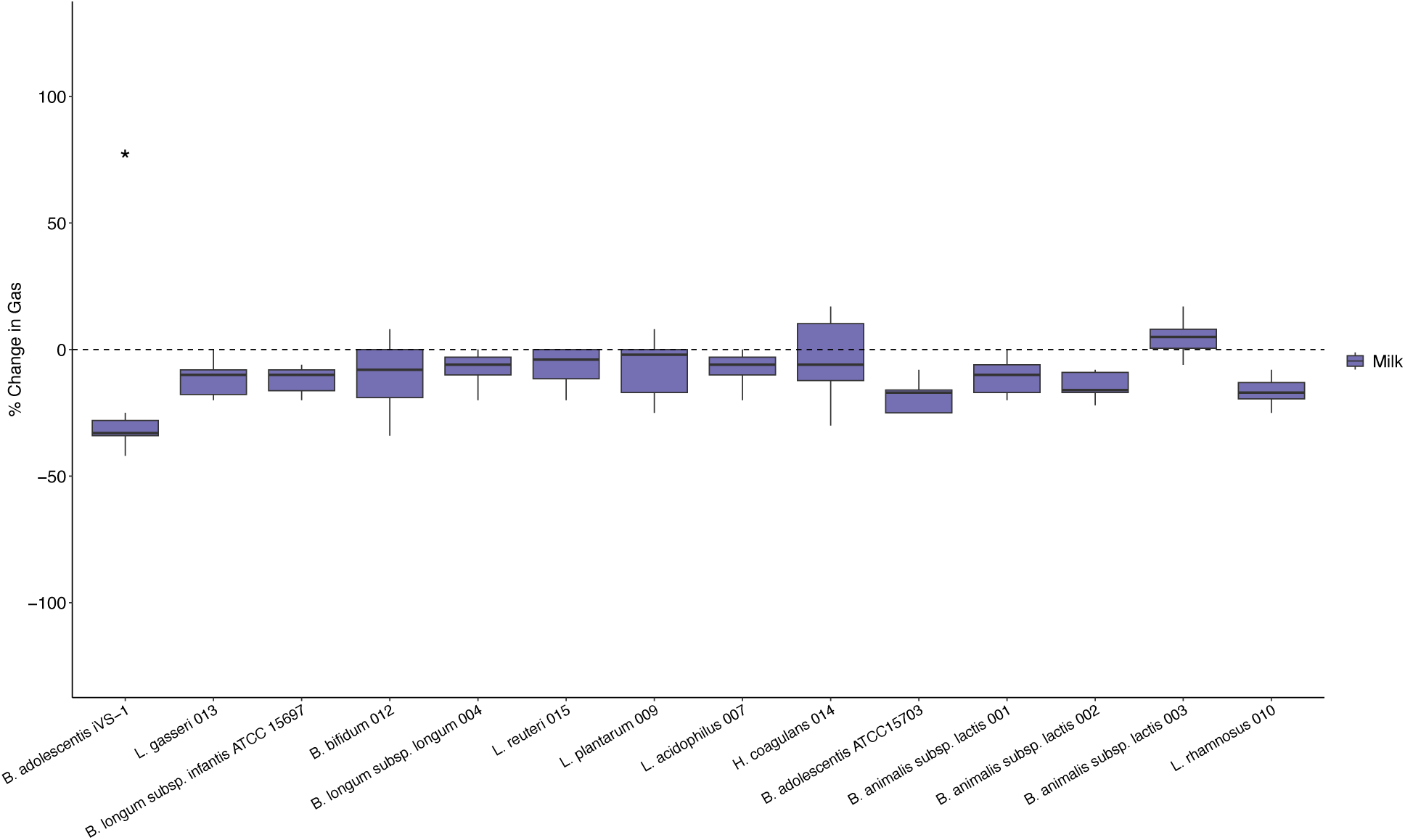
Gas reduction in milk. Change in gas produced by four fecal communities containing 5% by volume of 2% milk, relative to communities with no probiotic added. *p < 0.05 by Mann-Whitney-Wilcoxon test. See Supplementary Table 1 for data and p-values. Box plots show the interquartile range (IQR; boxes), median (line), and 1.5 IQR (whiskers).

**Supplementary Figure 3.**
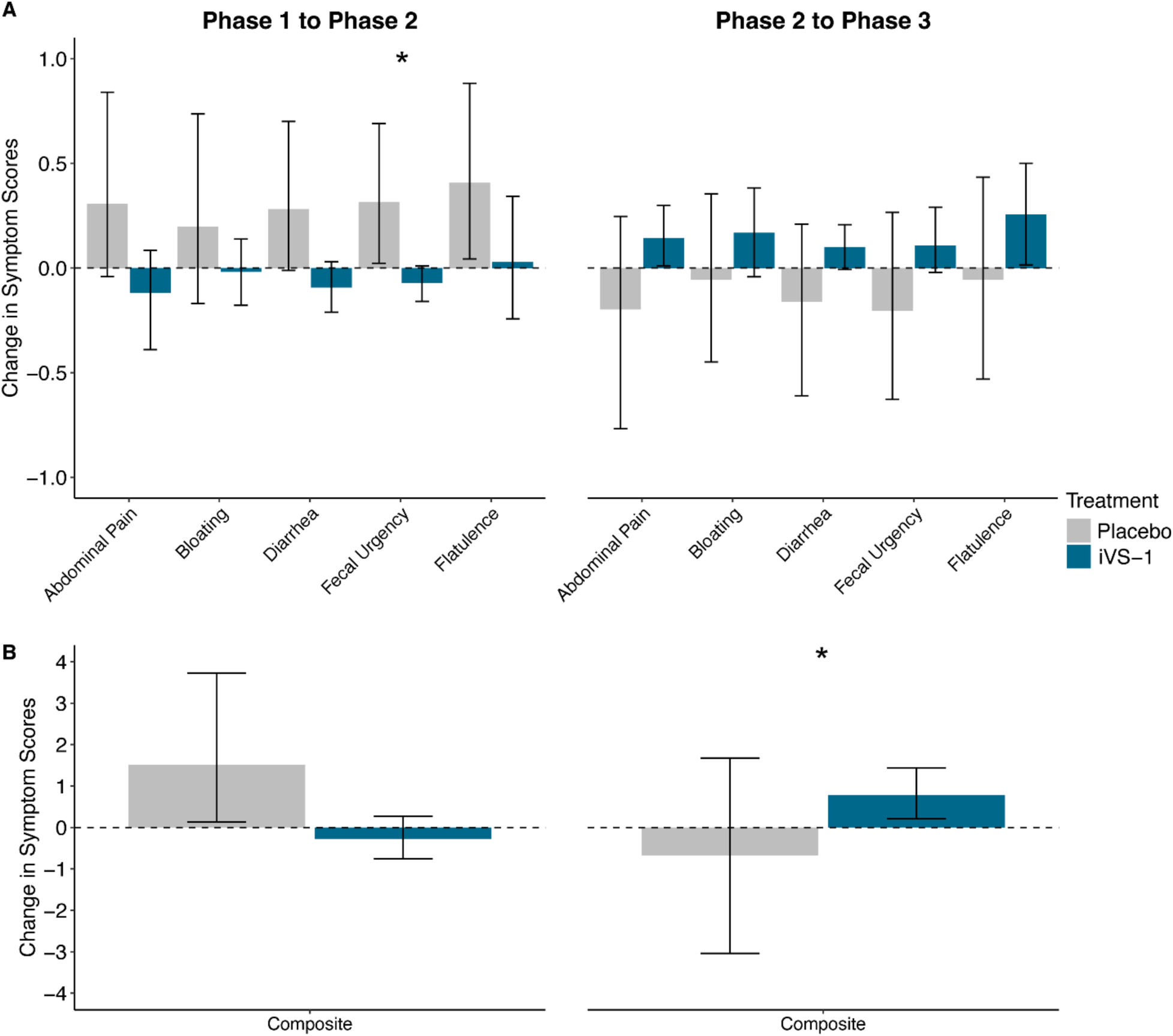
Change in patient symptom scores between study phases. A. Change in mean daily symptom scores for subjects by treatment time period (Phase 1 to Phase 2 and Phase 2 to Phase 3). B. Change in the composite (sum) of all mean daily symptom scores. n = 21 (Phase 1 to 2), 20 (Phase 2 to 3). *p < 0.05 by Mann-Whitney-Wilcoxon test. See Supplementary Table 1 for changes in means and p-values. Bars extend to the mean and whiskers span the 95% confidence interval.

**Supplementary Figure 4.**
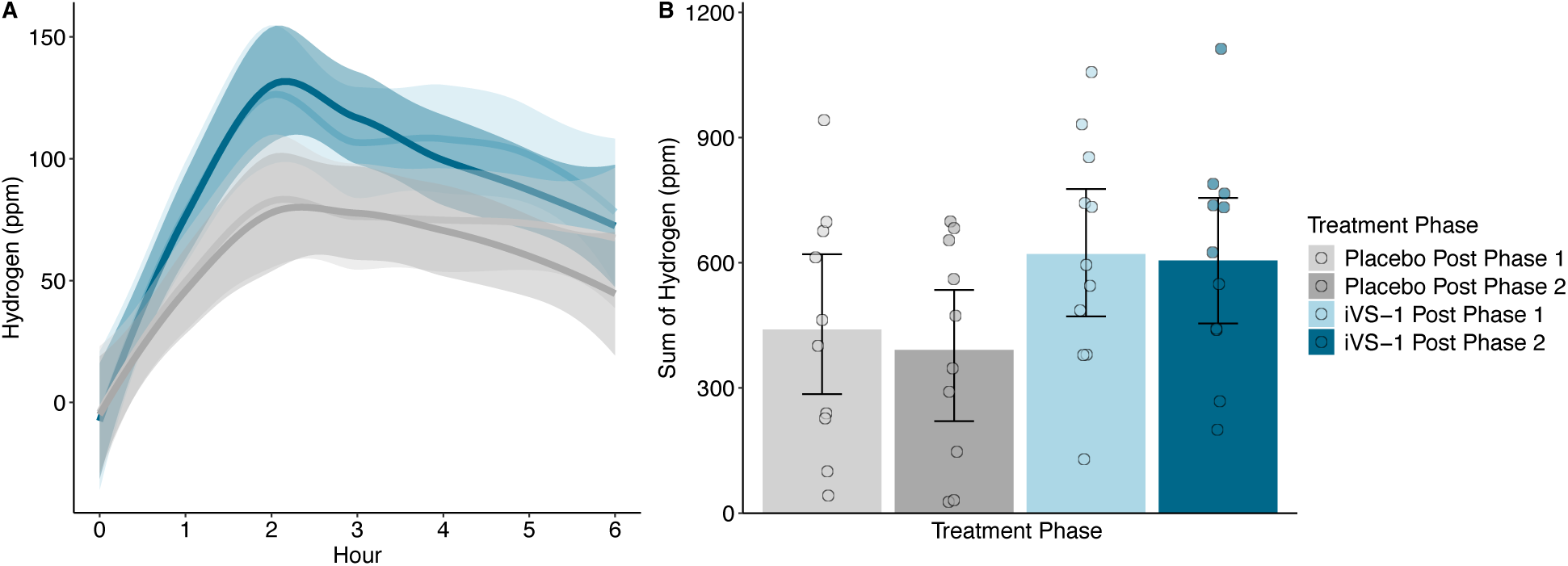
Subject breath hydrogen following lactose consumption. A. Breath hydrogen over time. Shaded regions represent 95% confidence intervals. B. Cumulative breath hydrogen from seven measurements over six hours. Each subjects’ data was normalized for background hydrogen production at time zero. Bars extend to the mean and whiskers span the 95% confidence interval. n = 21 subjects and all comparisons p > 0.05 by Generalized Estimating Equations modeling (A) and Mann-Whitney-Wilcoxon or Wilcoxon signed-rank testing (B). Post-Phase 1 occurred between Phase 1 (before treatment) and Phase 2 (during treatment). Post-Phase 2 occurred between Phase 2 (during treatment) and Phase 3 (after treatment). See Supplementary Table 1 for data and p-values.

**Supplementary Figure 5.**
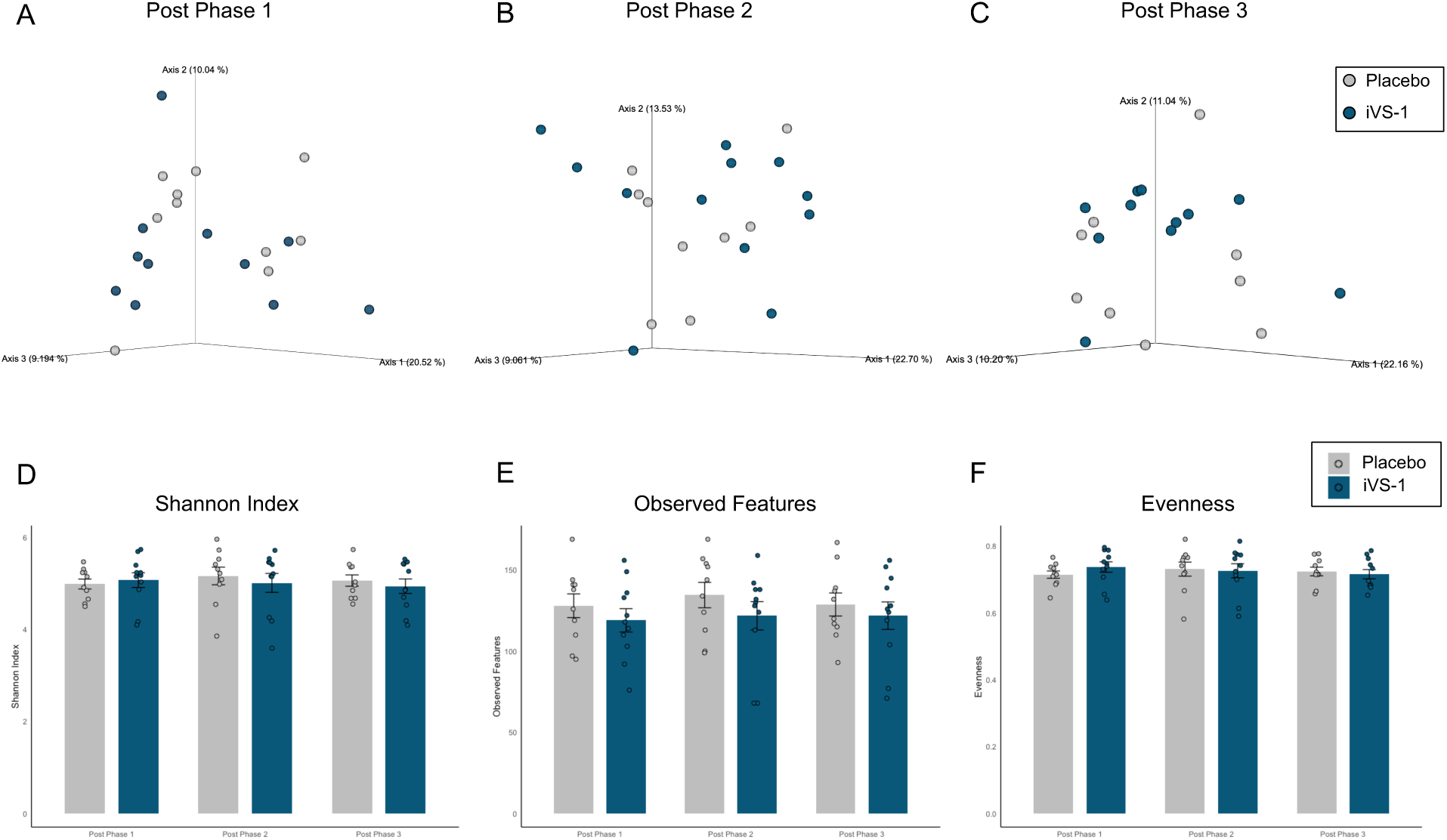
Alpha and beta diversity of iVS-1 and placebo groups. Principal coordinate analysis of unweighted UniFrac distances at post-Phase 1, 2, and 3 (A-C). Alpha diversity assessed by Shannon indices (D), observed features (E), and evenness (F). All p > 0.05 by pairwise PERMANOVA (A-C) and Mann-Whitney-Wilcoxon test (D-F).

**Supplementary Figure 6.**
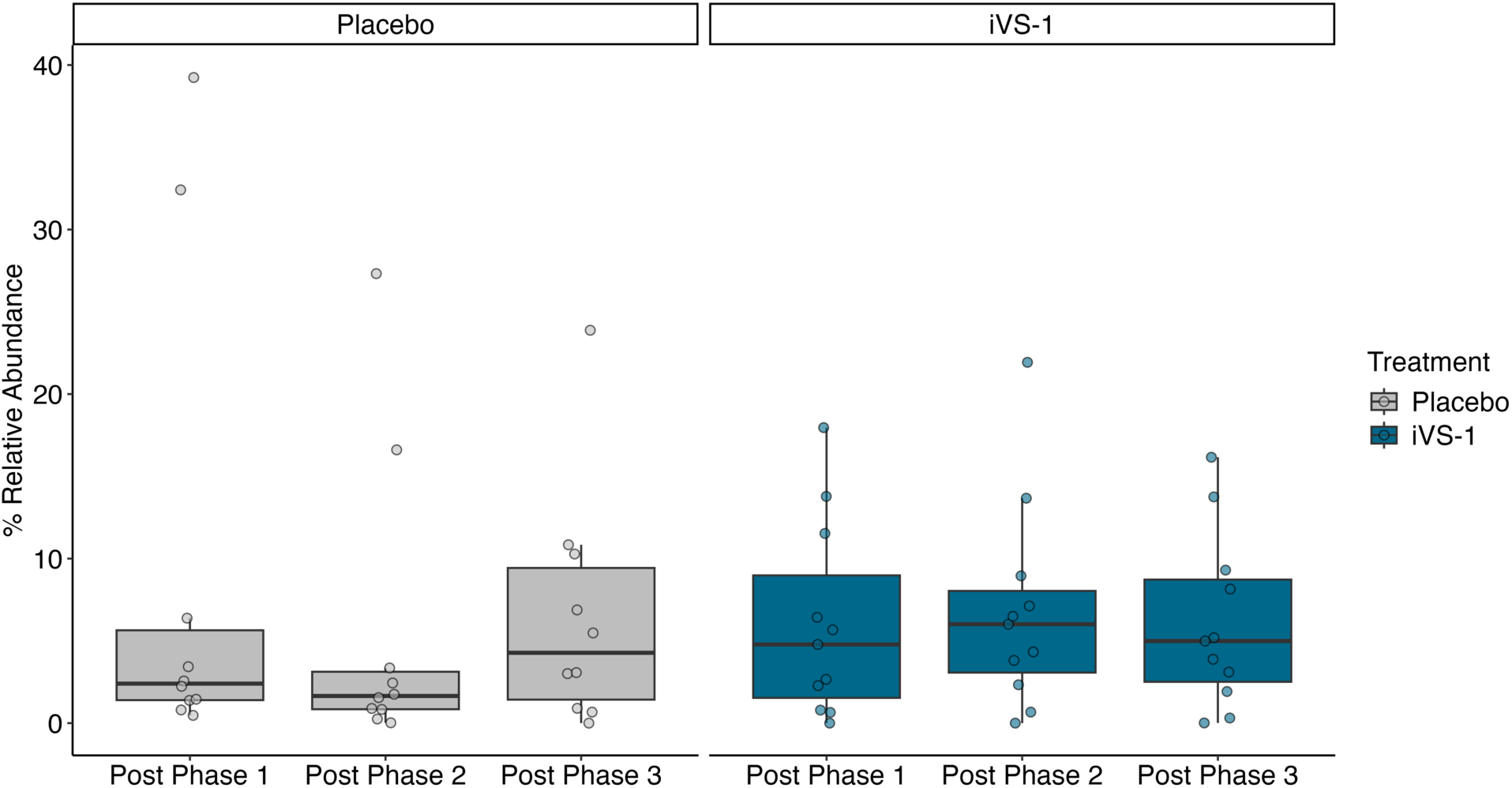
*Bifidobacterium* 16S rRNA gene relative abundance by post Phase. The relative abundance of *Bifidobacterium* 16S rRNA gene amplicon sequence variants was examined between iVS-1 (n = 11) and placebo (n = 10) groups post Phase 1, 2, and 3, and between phases for each treatment group. All p > 0.05 by Mann-Whitney-Wilcoxon test. See Supplementary Table 1 for data and p-values. Box plots show interquartile range (IQR; boxes), median (line), and 1.5 IQR (whiskers).

**Supplementary Figure 7.**
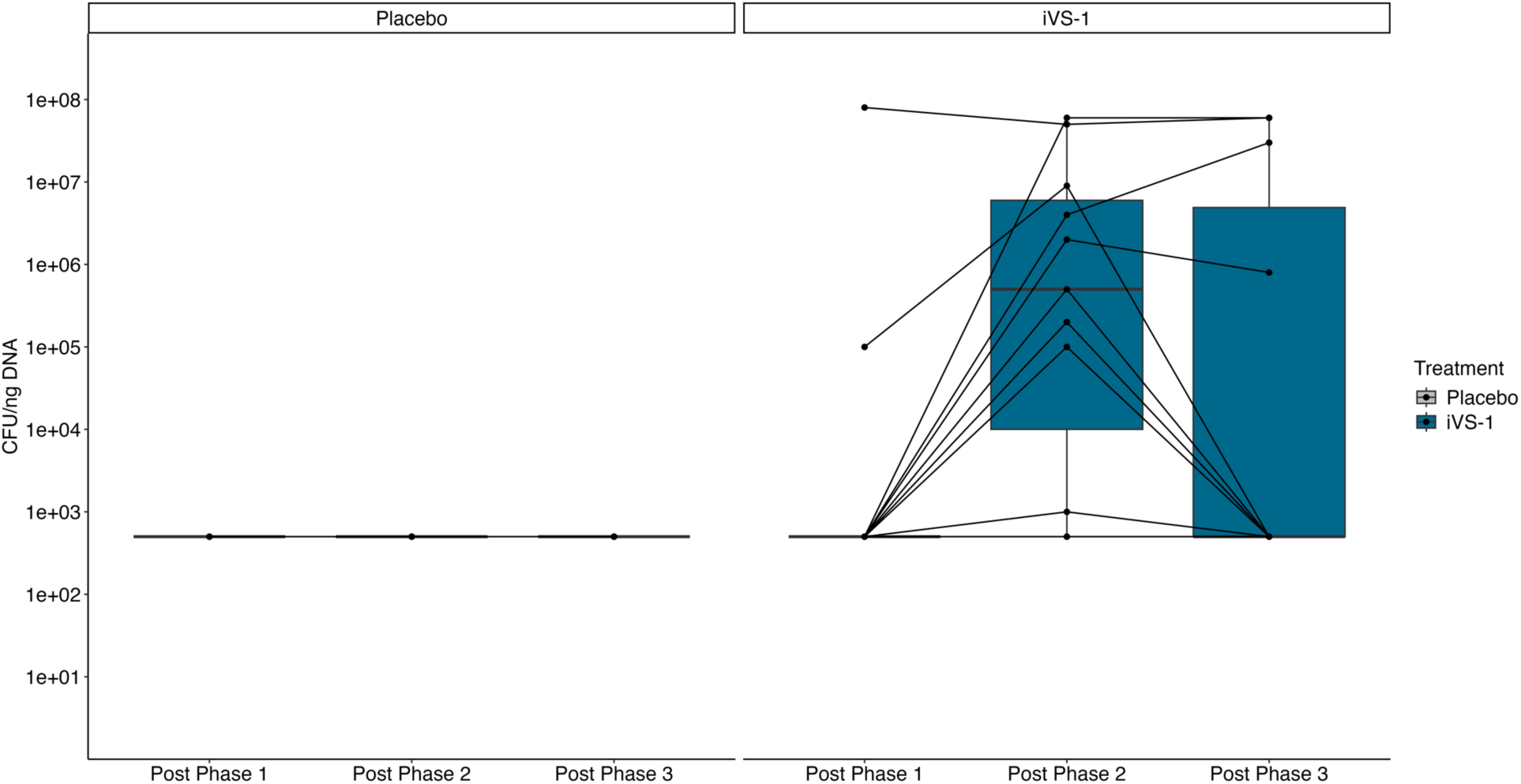
Abundance of iVS-1 in fecal samples. Using qPCR, the abundance of iVS-1 was determined following all three study phases. Points from individual subjects are connected by lines. n = 21. *p = 0.045 and ***p = 0.0005 by Mann-Whitney-Wilcoxon test. See Supplementary Table 1 for means and p-values. Box plots show interquartile range (IQR; boxes), median (line), and 1.5 IQR (whiskers).

**Supplementary Figure 8.**
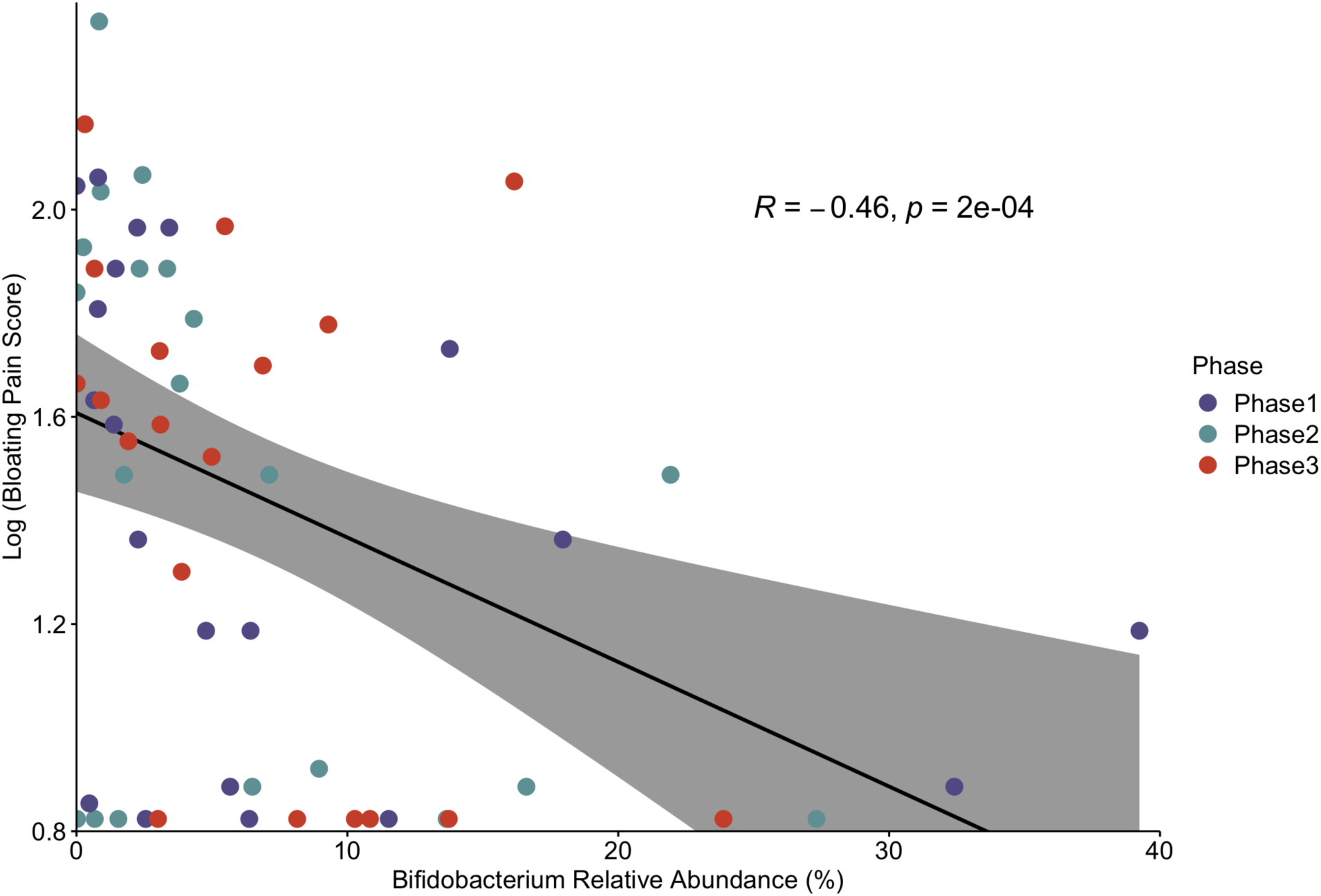
Correlation of *Bifidobacterium* relative abundance and bloating scores. Mean daily bloating scores were multiplied by 100 and log transformed. Subjects with zero symptoms were calculated with the lowest non-zero values to allow logarithmic scaling. Points are mean daily scores for individuals (n = 20) at each phase of the study (colors) matched with the relative abundance of *Bifidobacterium* 16S rRNA genes at the end of that phase. R = -0.46 (p = 0.0002) by Spearman correlation.

